# Applicability Area: A novel utility-based approach for evaluating predictive models, beyond discrimination

**DOI:** 10.1101/2023.07.06.23292124

**Authors:** Star Liu, Shixiong Wei, Harold P. Lehmann

## Abstract

Translating prediction models into practice and supporting clinicians’ decision-making demand demonstration of clinical value. Existing approaches to evaluating machine learning models emphasize discriminatory power, which is only a part of the medical decision problem. We propose the Applicability Area (ApAr), a decision-analytic utility-based approach to evaluating predictive models that communicate the range of prior probability and test cutoffs for which the model has positive utility; larger ApArs suggest a broader potential use of the model. We assess ApAr with simulated datasets and with three published medical datasets. ApAr adds value beyond the typical area under the receiver operating characteristic curve (AUROC) metric analysis. As an example, in the diabetes dataset, the top model by ApAr was ranked as the 23^rd^ best model by AUROC. Decision makers looking to adopt and implement models can leverage ApArs to assess if the local range of priors and utilities is within the respective ApArs.

## Introduction

Clinical predictive models are increasingly the focus of research, implementation, and digital health entrepreneurship^1,2^. Beyond calls for evaluation, in general^3^, leaders in machine learning (ML) in medicine call for considering “clinically acceptable performance characteristics for the targeted application”^4^. The reason for this call is that current methods do not incorporate the notion of “clinical acceptance” either in the derivation of the model or in its evaluation.

The decision faced by an institution considering a statistical or ML model (SMLM) is whether their patients would benefit from the model. From this perspective, the SMLM is a test, and the question is whether the utility of the population is maximized, at the prevalence of the disease they are treating, using this test. As such, the decision would be governed by a measure of “clinical acceptance”, which would indicate the fit of SMLM for use or otherwise. To establish “clinical acceptance,” proper utility tradeoffs should be articulated in the model construction and the model evaluation, coherent with the medical decisions faced by an institution^5^.

Regarding model constructions, at the heart of the derivation of any SMLM is a loss function that the learning algorithm uses to judge whether the model being developed is “close” to the training data^6^. These loss functions pit false positives (FPs) against false negatives (FNs) and, almost always, implement the assumption that those costs are the same^7,8^. While such symmetry has nice statistical properties, it is a false, and possibly dangerous, representation of the clinical context. For instance, treating 1 patient with antibiotics for a deadly infection they did not have is much more acceptable than having a patient with such an infection die because they were not treated, and even 50 patients unnecessarily treated might be acceptable^9^. One aspect of “clinically acceptable,” then, is to honestly elicit and incorporate that asymmetry in the SMLM creation process. We presume that the deciding clinician can make such an assessment and that individual patients may have different assessments.

Regarding model evaluation, the current standard evaluation of an SMLM is its discriminatory power, as assessed by the area under the receiver operating characteristic (AUROC) curve. The AUC also presumes that the cost of FPs and FNs is the same^10,11^. Furthermore, an AUC does not account for the disease prevalence in the population precisely because it evaluates the performance of a cutoff within each group (diseased and non-disease). However, the performance of a test, which role an SMLM plays in clinical practice, does depend on prevalence. Furthermore, each patient has a unique set of risk factors at the point of care (demographics, medical history, social determinants of health, etc.), which yield a different prior probability of disease; thus, any local population of patients contains a range of priors collectively rather than a single number. Hence, a “clinically acceptable” measure should account for ranges of priors as well as ranges of preferences.

Decision Curve Analysis (DCA) is one popular approach that integrates preferences into model evaluation^12,13^. DCA evaluates the model by calculating the net benefit over ranges of probability thresholds, keeping the prevalence constant. This constant prevalence does not account for the variability in prevalence within a site, let alone across different sites, where the SMLM is poised to be implemented. We present here a novel approach consistent with the SMLM implementation context, accounting for ranges of preferences and prevalence.

In this paper, we demonstrate that decision analysis provides the necessary theory to address model evaluation, deriving a novel measure called Applicability Area and demonstrating the implications of this measure, which turn out to upend several results based on AUCs. The objective of this paper is to illustrate the novel approach to model evaluation and compare it against the AUC traditional model performance metric. Additionally, we demonstrate the value of the novel approach with simulated data and with three medical datasets.

The rest of the paper is as follows: First, we present the decision analytic setup and the derivation of our measure. The following section presents the methods employed to demonstrate the use of our measure, including both simulated and real-world medical datasets. Then, we present the results and discuss the findings comparing AUC and our approach. Lastly, we compare the approach against DCA, a popular alternative that shares a similar goal.

### Decision Analytic Setup

A proper decision analysis entails making the best decision based only on information available at the time of making the decision, and the “best” decision is the one that maximizes the decision maker’s overall utility^14^. We demonstrate this approach using The Kassirer–Pauker formulation of threshold-based decision-making^15^. Although these decisions affect patients, who should be the decision maker for a single decision, in our model, we take the perspective of a provider contemplating applying the SMLM to their population of patients, with the valuation of outcomes based on patients’ expected utility (preference valuations under uncertainty^16-21^). In a simple decision model, either all patients are treated, or all patients are not treated, resulting in 4 health states: 1a) Treated and Disease (RxD), 1b) Treated and No Disease (RxNoD), 2a) Not treated, Disease (NoRxD), and 2b) Not Treated, No Disease (NoRxNoD) (the top two branches in Figure 1a). Of the 4 states, NoRxD is the worst, and NoRxNoD is the best. Utilities^16-21^ are assigned by the decision-maker to each state. The utility of NoRxD (uNoRxD) is usually 0 and that of NoRxNoD (uNoRxNoD), is usually 1; however, the recommendations of the model are the same under a linear transformation of the utilities.

**Figure 1.**
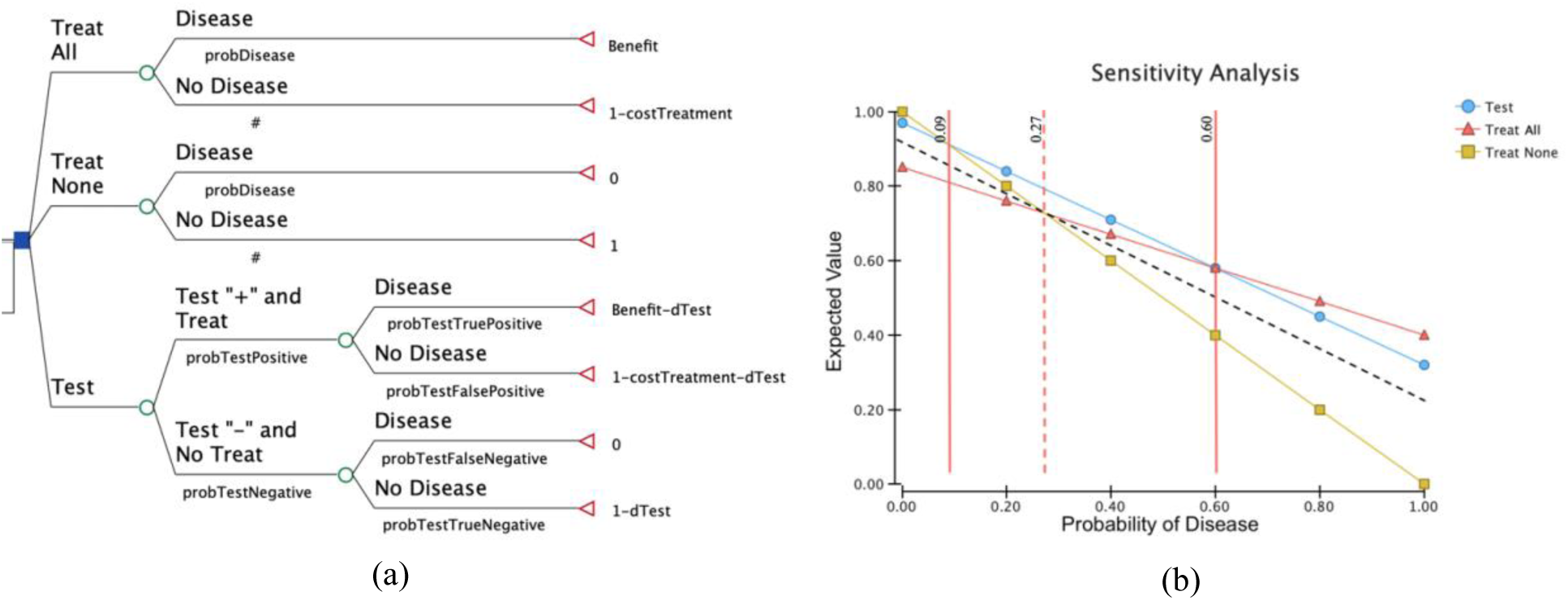
Kassirer–Pauker framework. (a) Alternatives of Treat All, Treat None, Test. Parameters are the probability, pDisease (probability of disease) and the utilities, Benefit, and cTreatment (cost (burden) of treatment) (b) Sensitivity analysis for pDisease, where the model was recalculated for several values across all possible values of pDisease (0 to 1). These calculations result in 3 thresholds, across which, the optimal strategy changes: for pDisease < the Treat None/Test threshold, “Treat None” is the optimal strategy; for pDisease > Test/Treat All threshold, “Treat All” is the optimal strategy; and for pDisease between the two thresholds, “Test” (that is, use the model), is optimal. The values of the thresholds are functions of the cost asymmetry and the sensitivity and specificity of the test. Since each cutoff on the ROC generates a different sensitivity and specificity, there will be a different “Test” line (and therefore, a set of thresholds) for each cutoff on the ROC curve. For some cutoffs, those points may result in a line that crosses the threshold vertical, *pDisease* = *p*^*\**^ below where the lines for Treat None and Treat All cross. For those cutoffs, the model (test) is not useful. dTest is the cost (burden, disutility) of the test.

In a simple decision tree (Treat All or Treat None) one calculates the expected utility of each option. In this case, the key probability is *p* (disease probability) and is the same for both subtrees. Expected utility is calculated as the sum of the product of a subtree’s probability and its utility. In this case EU(Treat All) = *p* × uRxD + (1–*p*) × uRxNoD, and EU(Treat None) = *p* × uNoRxD + (1–*p*) × uNoRxNoD. The model then reports which expected utility is higher. Since higher utility is preferred, the model then “recommends” the decision maker take the action with the higher expected utility. There is a probability, *p*\*, where the expected utilities are the same. This probability, *p*\*, is derived by setting the two expected utilities as equal.

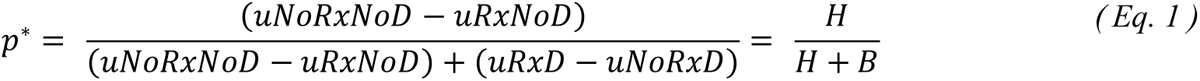

uNoRxNoD – uRxNoD is the difference to a non-diseased patient between not being treated and being treated (unnecessarily), so is labeled, H, the harm of treatment (to the non-diseased patient). The quantity uRxD –uNoRxD is the difference to a diseased patient between being treated and not, and so is labeled, B, the benefit of treatment (to the diseased patient). The ratio, B/H, will provide us with the asymmetry factor we need. For instance, 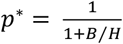. If harms were the same as benefits, the threshold would be 0.5. The “clinical context” is therefore represented by the prior and the asymmetric costs. This choice fits with the recurring recommendations that models need to be checked against local constraints^5,22-25^.

An SMLM is used clinically as a test. To accommodate this “test,” we need to add a third subtree to the initial model (see the bottom branch in Figure 1a). The utility formula is more complicated because it needs to consider 4 states: Test “positive” (resulting in true positives (TP) and false positives (FP)) and Test “negative” (resulting in true negatives (TN) and false negatives (FN)). The 4 test states correspond to the 4 health states (Test “+” Treated and Disease, Test “+” Treated and No Disease, Test “-” Not treated and Disease, and Test “-” Not Treated and No Disease). Thus, the benefit of the test is the difference between the utility of a TP and the utility of an FN (uTP – uFN), and the harm of the test is the difference between the utility of a TN and the utility of an FP (uTN – uFP). The probabilities are now different because Bayes’ Theorem provides the positive and negative predictive values (P(D+|Test +) and P(D–|Test –), respectively). One can add in the (dys)utility of the test itself (dTest).

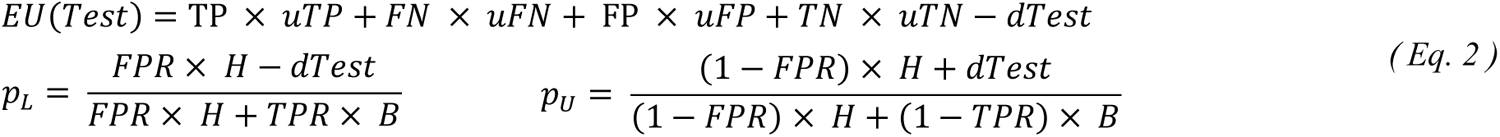

Figure 1b shows a sensitivity analysis with 2 thresholds: The lower one between Treat None and Test (pL) and the upper one, between Test and Treat All (pU). These thresholds are functions of sensitivity, specificity, and the utilities (H, B), and dTest. For a test to be useful, there must be two thresholds, which is equivalent to saying that the utility line for EU(Test) must cross the line *p*= *p*\* above where the lines for EU(Treat All) and EU(Treat None) cross. For a test to be useful, furthermore, the current prior must be within the bounds of the two thresholds.

#### Derivation of Applicability Area

The formula for the ideal cutoff of a test is well-known ^5,9^, 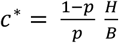. The test cutoff is different from the probability threshold *p*\*, treat all or treat none. *In an SMLM that produces a probability as its output, the test cutoff, which is the probability that separates a positive from a negative class, is the same as the posterior probability threshold*. This formula makes clear that the ideal cutoff for an SMLM depends on local priors (*p*) and asymmetric costs. While this formula tells us how to apply an SMLM, most developers want a metric to show that their model is better than other models and to show that their model works in a range of settings, that is both a range of priors and a range of posterior probability thresholds^26^.

We call the range of priors and posterior probability thresholds in which an SMLM is useful, its area of applicability. *A model’s applicability is the cumulative range of priors in which the overall utility of the model is greater than that of the alternative treat-none and treat-all strategies*. Outside this area, the decision maker knows what to do: either to treat or not. The applicability of a model incorporates the relative cost tradeoffs for each outcome and model discrimination (sensitivity and specificity).

Given a ROC, each cutoff, *c*, generates its own FPR(*c*) and TPR(*c*), and, therefore, its own Treat None/Test and Test/Treat All thresholds:

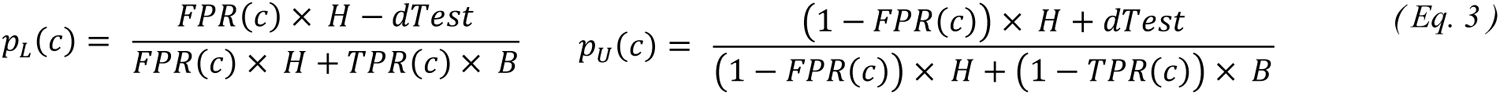

The thresholds are functions of the costs and benefits. There will be a minimum cutoff, *c*_min_, below which there is no lower threshold, and a maximum cutoff, *c*_max_, above which there is no upper threshold. Similarly, there is a minimum prior, *π*_min_, and a maximum prior, *π*_max_, the regions between which, the SMLM remains “useful.” Thus, the Applicability Area is formulated as Eq. 4:

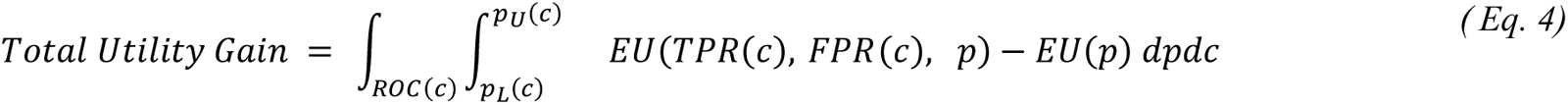

Total Utility Gain is calculated as a double integral, first, across all cutoffs of the ROC, then between the two thresholds, for a given cutoff, of the difference between the expected utility using the SMLM and the expected utility without the SMLM. To speed the calculation and to make ApArs more comparable across models, we simply give “credit” for positive utilities. In other words, the integrand would be 1 if the difference in utility between using and not using the SMLM is greater than 0, and 0 otherwise.

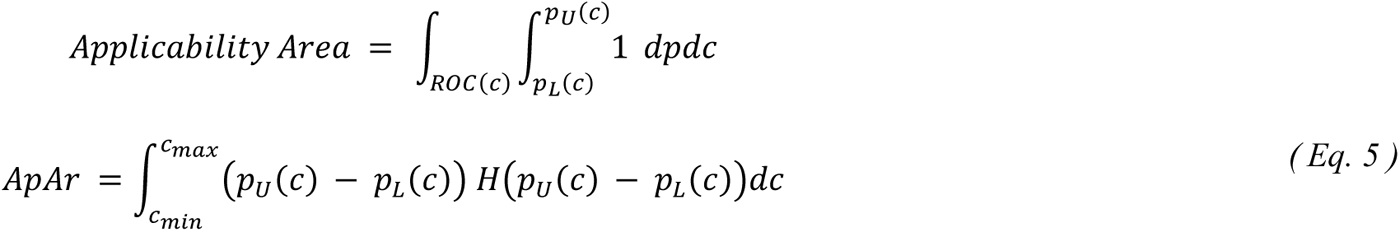

The criterion, *p*_*U*_(*c*) > *p*_*L*_(*c*), is equivalent to specifying that the differences, EU(model) – EU(Treat All) and EU(model) – EU(Treat None), are both positive. As we showed in Figure 1c and Eq. 3, the model is only useful when there are two test-related thresholds, i.e., *p*_*U*_(*c*) > *p*_*L*_(*c*). Thus, we integrate over only the region satisfying this condition. Figure 2 shows an example of a model’s applicability area. Since the maximum possible ApAr spanning priors and posterior probabilities is 1, the ApAr would be less than or equal to 1. The larger the ApAr, the model is applicable over wider ranges of priors and posterior probability thresholds.

**Figure 2.**
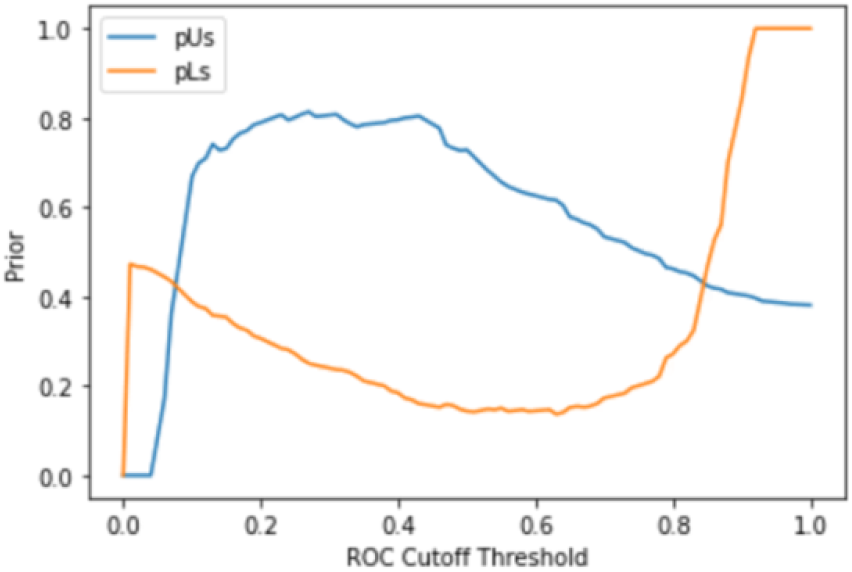
Example of a model’s applicability area, where *p*_*U*_(*c*) > *p*_*L*_(*c*). ApAr is the area bounded by the blue and orange curves when the blue curve is above the orange (in this case, between cutoffs of about .1 and .85).

A different set of utility curves such as the one shown in Figure 1b can be generated for each cutoff point on the ROC. As established earlier, each utility graph contains 2 thresholds (the testing threshold, pL, and the treatment threshold, pU). The applicability metric involves 1) calculating the range of priors from the two thresholds (*p*_*U*_(*c*) − *p*_*L*_(*c*)) and 2) integrating over the entire ROC to obtain the cumulative ranges of applicable priors (Eq. 5). Hereon, “applicable priors” refer to the range of priors under which the model will have positive utility or is preferred over the alternative strategies. For AUC, points closest to the upper left are generally presumed to suggest preferred cutoffs^11^. Indeed, in our results, probability thresholds closer to the upper left corner of the ROC have larger ApArs. However, such a high-AUC model may not be useful elsewhere along the ROC. Our approach considers all the ranges of priors for which the model is useful. It describes the model performance in terms of aggregated applicable priors under all cutoffs rather than a single cutoff in the upper-left corner. We eliminate the need to determine the prior beforehand, which can be difficult^16,27^. A model with an applicability area (ApAr) of zero indicates employing the model as a test for the probability of disease has no value compared to a treat-none or treat-all strategy. On the other hand, high applicability indicates that the model is useful as a test for the probability of disease over greater ranges of priors.

## Method

### Simulation

The simulation aimed to assess the circumstances in which the ApAr gives results different from the standard ROC AUC. After discussion with colleagues and a review of the literature^28^, we settled on 5 factors that might affect the measure’s performance: sample size, number of variables, class proportion, covariance, and class separation. In this fully factorial simulation, we varied cost asymmetry and the 5 factors. Cost asymmetry, which represents the cost ratio between a minority and a majority class, carried 4 discrete values (1, 2, 5, and 10). The 5 factors were specified as the following:

1. The sample size: 100, 5000, and 10,000 (small to large).
2. The number of variables: 2, 5, and 15 (few to many).
3. Covariance among the predictors: 0.05, 0.1, and 0.2 (low to mild).
4. Class proportion: 0.1%, 1%, 10%, and 30% (uncommon to frequent)^29^.
5. Class separation: 0.05, 0.1, and 0.2 (small to modest).

Scikit-learn library’s method, `make_classification`, was modified to admit covariance as an additional parameter^30^. Then, all 5 factors were fed into this modified method. A total of 1296 simulated datasets were generated.

### Statistical analysis

Each dataset was fitted to 5 SMLM: logistic regression (LR), xgBoost, decision trees (DT), random forest (RF), and support vector machine (SVM). Performance metrics were ROC AUC and ApAr. With a true negative as the best outcome and a false negative as the worst outcome, the relative values of uTN and uFN were 1 and 0 respectively. We assigned uTP a typical value of 0.8 to represent a relatively beneficial disease treatment outcome. We assigned the population disease prevalence to 20%. Note that this prevalence reflects the external real-world condition, while “class proportion” reflects the prevalence of the real-world condition in the dataset. uFP is dervied from B/H and B.

#### Medical Datasets

We also used three imbalanced medical datasets to evaluate our approach: the Pima Indians Diabetes (PID), Cervical Cancer Risk Factors (CC), and Chronic Kidney Disease Risk Factor datasets (CKD)^31-34^. The PID dataset was selected from a larger database created by the National Institute of Diabetes and Digestive and Kidney Diseases. It was created to develop diagnostic prediction models for diabetes. All patients in this dataset were females of Pima Indian heritage and over the age of 21. The predictors in this dataset include age, the number of pregnancies, glucose concentration, blood pressure, skin thickness, insulin level, Body Mass Index (BMI), and diabetes pedigree function score. The CC dataset originates from the University Hospital of Caracas in Venezuela. The dataset captures 858 patients and includes data on demographics, medical history, and lifestyle. Like other studies that used this dataset, we used the set of features from the study led by Fernandes et al.^32^. The CKD dataset, originating from the Apollo Hospitals in India, was collected over 2 months. The dataset captures 400 patients, and it includes patients’ age, comorbidity, diet, and laboratory test values such as red and white blood cell concentration.

### Statistical analysis

No missing values existed in the PID dataset. However, both the CC and CKD datasets contained missing values for some of the predictors. Without the facility to check for missingness not at random, we used K-nearest neighbor imputation with default hyperparameters (n = 5 and metric = Euclidean distance) from the scikit-learn library to impute missing data^35^. Categorical predictors in the CKD dataset were also recoded as numeric variables and rescaled.

Each dataset was fitted to 5 SMLM: LR, xgBoost, DT, RF, and SVM. The cost ratio between the minority and majority classes varied among 1, 2, 5, 10, 20, 50, 100, and including the inverse of the class proportions as a heuristic for the relative misclassification costs^8,36^. 40 different models were created. As in the simulations, the relative values of uTN and uFN were 1 and 0, and uTP had a value of 0.8. uFP is dervied from B/H and B. Three repeats of 10-fold cross-validation were used to evaluate the models. Model performance was compared using the AUC and the ApAr. For each medical dataset, we compared the top ten models ranked by AUC along with the top 10 models ranked by the ApAr. All simulation programming and computations were performed using Python 3.7 and the scikit-learn ML library.

The code for Applicability Area is available on GitHub at: https://github.com/StarLiu1/ApplicabilityArea-ApAr

## Results

### Simulation results

From the simulation, 6480 classification models were generated from the 1296 datasets. **Figure 3**. plots each model’s AUC and ApAr to assess the level of agreement between the two metrics. While in general, ApAr increased as AUC increased, there was a lack of agreement between the AUC and the ApAr, as some of the models with the highest AUCs had the lowest ApArs, with agreement worsening as the asymmetry in costs increased. Conversely, several models with different AUCs had the same ApAr. Breakdown by each SMLM shared the same pattern.

**Figure 3.**
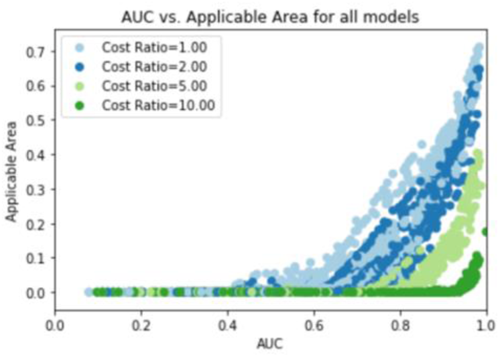
AUCs vs. applicability areas for each model as a point. Each of the four curves corresponds to a different B/H cost asymmetry.

We obtained the test cutoff that gave the largest range of useful priors for each model and compared the model’s AUC and ApAr (Figure 3). Models with the largest range of applicable priors had the highest AUCs. However, after incorporating tradeoffs (B, H), some of the models in the upper-left corner had the lowest ApAr, despite their high AUCs (Figure 4).

**Figure 4.**
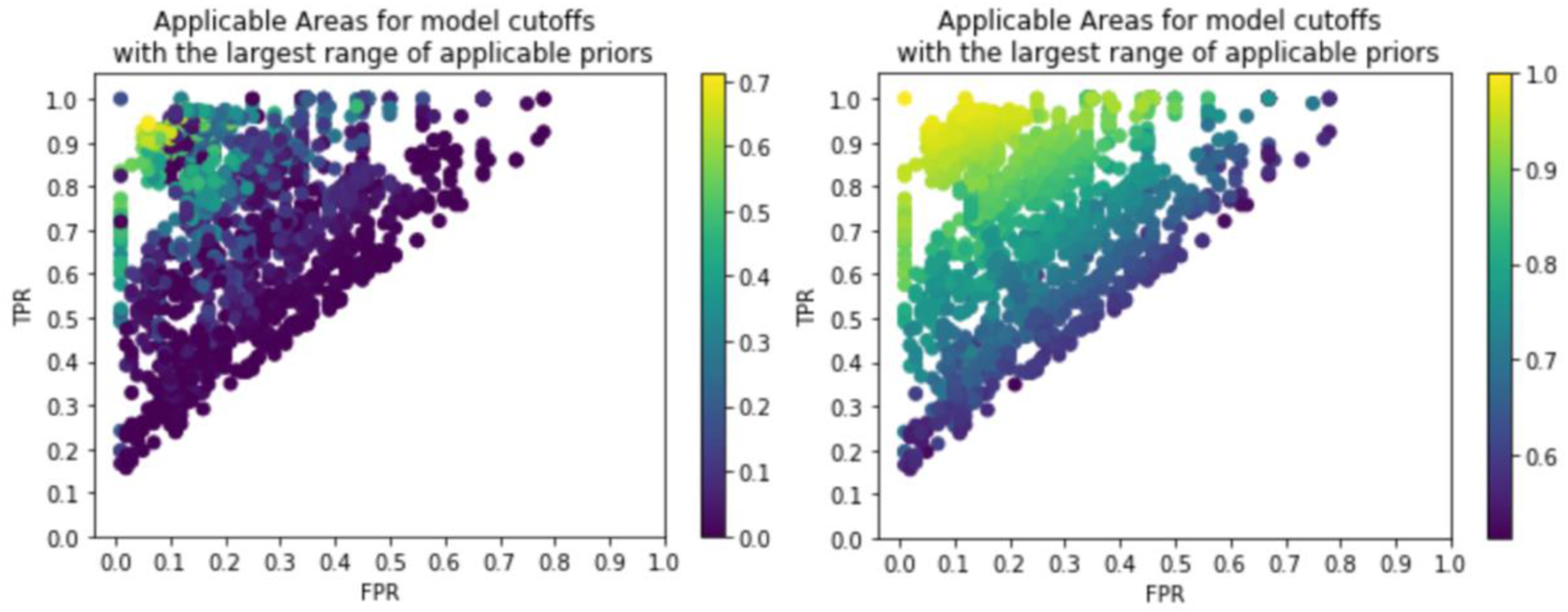
Comparing models’ applicability area (left) and AUC (right) at the cutoff with the largest range of applicable priors. The color indicates each model’s ApAr on the left graph and the AUC on the right graph.

### Medical Datasets

As shown in Table 1, there lacked agreement in the PID dataset models’ AUC and ApAr. Only 4 of the top 10 models were considered useful models by the ApAr metric. The model with the highest AUC (0.835) had the second-highest ApAr (0.252), and another model with the same AUC had the 6th-highest ApAr (0.178). In contrast, *the best model ranked by ApAr had the 23rd-highest AUC*. Similarly, we observe the same pattern in the CC dataset models’ AUC and ApAr (Table 1b). Despite universally high AUCs (> 0.94), only 4 out of the top 10 models were considered useful models by the ApAr metric. The model with the highest AUC (0.964) had the 16th-highest ApAr (0.010). The model with the second-highest AUC had the third-highest ApAr (0.350). In contrast, the model with the highest ApAr had the 15th-highest AUC (0.945). Lastly, we observe the same pattern in the CKD dataset models (Table 1c). Although the models had near-perfect AUCs (0.998-1.000), not all the models with perfect AUCs were considered useful by the ApAr metric. Even among perfect discriminators, ApAr ranged from 0.553 to 0.810.

**Table 1.** Comparing the models’ area under the curve (AUC) and applicability area (ApAr) using 3 datasets.*

| (a) Using the Pima Indians Diabetes dataset |  |  |  |  |  |  |  |  |  |
| --- | --- | --- | --- | --- | --- | --- | --- | --- | --- |
| Ordered by AUC |  |  |  |  | Ordered by ApAr |  |  |  |  |
| Model | Classifier | B/H | AUC | ApAr | Model | Classifier | B/H | AUC | ApAr |
| 0 | LR | 1 | 0.835 | 0.252 | 2 | xgBoost | 1 | 0.795 | 0.286 |
| 5 | LR | 2 | 0.835 | 0.178 | 0 | LR | 1 | 0.835 | 0.252 |
| 15 | LR | 5 | 0.835 | 0.025 | 4 | SVM | 1 | 0.824 | 0.227 |
| 20 | LR | 10 | 0.835 | 0 | 3 | RF | 1 | 0.822 | 0.217 |
| 10 | LR | 2.87 | 0.835 | 0.114 | 7 | xgBoost | 2 | 0.793 | 0.207 |
| 25 | LR | 20 | 0.834 | 0 | 5 | LR | 2 | 0.835 | 0.178 |
| 30 | LR | 50 | 0.833 | 0 | 9 | SVM | 2 | 0.827 | 0.160 |
| 35 | LR | 100 | 0.832 | 0 | 8 | RF | 2 | 0.827 | 0.148 |
| 8 | RF | 2 | 0.827 | 0.148 | 12 | xgBoost | 2.87 | 0.792 | 0.135 |
| 9 | SVM | 2 | 0.827 | 0.160 | 1 | DT | 2.87 | 0.665 | 0.116 |
| (b) Using the CC dataset |  |  |  |  |  |  |  |  |  |
| Model | Classifier | B/H | AUC | ApAr | Model | Classifier | B/H | AUC | ApAr |
| 23 | RF | 10 | 0.964 | 0.010 | 2 | xgBoost | 1 | 0.945 | 0.420 |
| 3 | RF | 1 | 0.961 | 0.350 | 7 | xgBoost | 2 | 0.948 | 0.363 |
| 28 | RF | 20 | 0.961 | 0 | 3 | RF | 1 | 0.961 | 0.350 |
| 13 | RF | 15.6 | 0.961 | 0 | 1 | DT | 1 | 0.772 | 0.332 |
| 33 | RF | 50 | 0.958 | 0 | 0 | LR | 1 | 0.921 | 0.255 |
| 8 | RF | 2 | 0.955 | 0.249 | 8 | RF | 2 | 0.955 | 0.249 |
| 38 | RF | 100 | 0.953 | 0 | 5 | LR | 2 | 0.919 | 0.235 |
| 18 | RF | 5 | 0.952 | 0.088 | 6 | DT | 2 | 0.768 | 0.212 |
| 17 | xgBoost | 5 | 0.949 | 0.161 | 9 | SVM | 2 | 0.914 | 0.201 |
| 7 | xgBoost | 2 | 0.948 | 0.363 | 4 | SVM | 1 | 0.875 | 0.180 |
| (c) Using the CKD dataset |  |  |  |  |  |  |  |  |  |
| Model | Classifier | B/H | AUC | ApAr | Model | Classifier | B/H | AUC | ApAr |
| 39 | SVM | 100 | 1.000 | 0 | 3 | RF | 1 | 1.000 | 0.810 |
| 29 | SVM | 20 | 1.000 | 0 | 4 | SVM | 1 | 0.999 | 0.776 |
| 3 | RF | 1 | 1.000 | 0.810 | 8 | RF | 2 | 1.000 | 0.765 |
| 8 | RF | 2 | 1.000 | 0.765 | 2 | xgBoost | 1 | 0.999 | 0.755 |
| 13 | RF | 2.67 | 1.000 | 0.726 | 0 | LR | 1 | 0.998 | 0.742 |
| 24 | SVM | 10 | 1.000 | 0.256 | 9 | SVM | 2 | 0.999 | 0.731 |
| 19 | SVM | 5 | 1.000 | 0.553 | 13 | RF | 2.67 | 1.000 | 0.726 |
| 34 | SVM | 50 | 1.000 | 0 | 5 | LR | 2 | 0.998 | 0.720 |
| 32 | xgBoost | 50 | 0.999 | 0 | 7 | xgBoost | 2 | 0.999 | 0.712 |
| 37 | xgBoost | 100 | 0.999 | 0 | 14 | SVM | 2.67 | 0.999 | 0.693 |

\* Green highlights indicate the same set of models that appeared in the top ten ordered by each metric. B/H = Benefit/Harm; LR = Logistic Regression; DT = Decision Trees; RF = Random Forest; SVM = Support Vector Machine

## Discussion

In this paper, we presented the motivation, derivation, and performance of a novel metric, ApAr, for evaluating predictive-model performance. In contrast to the traditional AUROC, it reports the range of priors for which the SMLM might be useful and bakes asymmetric cost into its formulation. We applied the approach to a simulated dataset as well as 3 medical datasets.

Our findings from the simulated dataset showed that, at the same Benefit/Harm tradeoff, models with the same AUROC did not have the same ApAr. Despite having the same AUC, the two models would have different tradeoffs in sensitivity and specificity in different regions on the ROC. The traditional procedure of model selection almost always selects the model with the highest AUC; however, in our simulations, this selection did not equate with the most applicable/practical/useful model overall. By representing asymmetric tradeoffs in model evaluation, the ApAr highlights models that are useful over ranges of priors (disease prevalence) and test cutoffs (posterior probabilities, in the case of SMLMs) thresholds. The ApAr approach evaluates model performance under contexts that are consistent with the decision-making.

We had similar findings from three medical datasets. Some of the models with the highest AUC were the least useful models given certain tradeoffs and priors. Conversely, some of the models with the highest ApArs had the lowest AUCs. Under the typical selection procedure, only the best-performing model would be selected. Thus, models with the highest ApAr would not have been selected if AUC were used as the metric, overlooking models that would have been more useful under certain ranges of local prevalence and posterior probability thresholds. As evidenced by models generated from the CKD dataset, even models with near-perfect AUCs (0.999-1.000) could lack applicable contexts given certain asymmetric tradeoffs and priors. In other words, choosing models based on discrimination only without considering the decision-analytic setup could lead to harmful decisions.

Models with the same ApAr would differ in either the range of applicable priors or in the range of posterior probability thresholds. For example, the top two models with the highest ApAr using the PID datasets had different AUCs (0.793 vs. 0.835) and near identical ApAr (0.024 and 0.025). However, if we plot the ApAr, we see that the models were applicable over different ranges of priors and posterior probability thresholds (Figure 5). As the AUC increases, the ApAr becomes more rectangular like that of Figure 6. Higher AUC would equate with greater separation in the posterior probabilities. Thus, a model with a high AUC would be applicable over ranges of prior along the entire ROC regardless of the classification cutoff. Two models with the same AUCs and ApArs could have different ranges of applicable priors and posterior probability thresholds. A detailed breakdown of the two models is needed before selecting the most appropriate one. Finally, the best cutoff depends on the *c*^∗^ formula^9^.

**Figure 5.**
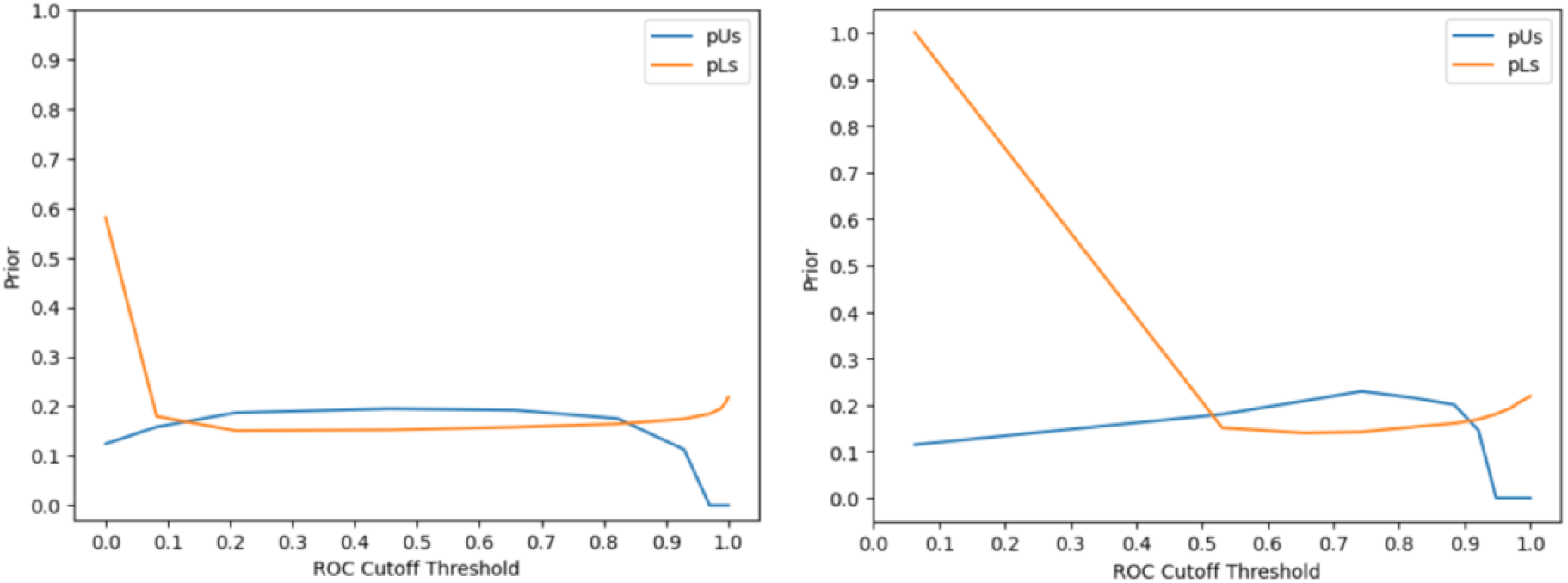
Comparing the Applicability Areas of models with nearly the same ApAr (Left: 0.024; Right: 0.025)

Another decision-analytic-based approach, DCA, has gained traction in recent years.^12,13^ DCA has been a popular method that attempts at evaluating models by incorporating harms and benefits. Like the DCA, we show that the AUC can be misleading beyond discrimination and can even lead to choosing an underperforming model. The major difference is that DCA keeps the prior constant and varies the threshold, *p*\*, implicitly being B/H; ApAr keeps a population-level cost asymmetry B/H constant and reports the ranges of priors and thresholds for which the model is useful. These ranges account for individual patients having different pre-test probabilities and different assessments of cost asymmetry than the policy-making clinician. The next step in developing the ApAr is showing the ranges of population-level B/H assessments for which a model is useful.

APLUS is a novel framework that simulates care management workflows to assess the usefulness of ML models for deployment and integration^37^. It models nearly any workflow by defining workflows as sets of discrete states and transitions. It models patients’ trajectories and allows for utility tradeoffs and cost parameter specifications. Lastly, APLUS also enables human-readable outputs. However, the core limitation of APLUS is that it requires significant effort in defining the workflow steps and in involving multiple stakeholders to accurately represent the workflows, tradeoffs, and uncertainties.

We might suggest the ApAr, by reporting ranges of priors and thresholds, helps decide whether to consider a model at all. DCA helps decide whether local thresholds imply net benefit. APLUS helps with the final local adapatation^22,38^.

### Strengths

The advantage of the novel applicability approach is several folds. First, the utility-based approach clearly articulates the tradeoffs behind a decision that the SMLM model is designed for. The approach ensures a transparent cost structure consistent with the represented decision problem. Second, the approach redefines the usefulness or impact of an SMLM model based on utilities rather than heuristics and enables meaningful comparisons across models. A useful model under the applicability approach is one associated with practical and cumulative ranges of priors. Third, considering the ranges of applicable priors eliminates the need to choose a prior before conducting the analysis, which has long been a challenge^25^. Fourth, the ApAr approach is SMLM agnostic because it requires only the ROC, which can be generated from any prediction model. Lastly, the utility-based approach directly addresses two questions central to adopting an SMLM: 1) is the model useful at all? 2) when is the model useful? (as priors and utilities vary locally). Institutions looking to implement SMLM in clinical decision support tools can use this approach to compare models and select the one that best fits the institution’s take on cost asymmetry and the implicit priors in the target population.

### Limitations

There remain several limitations to our approach. First, model developers have pointed out the challenge of quantifying the tradeoffs and have avoided explicit representations. Our approach was not intended to provide a solution, but rather highlight the need to clearly articulate the tradeoffs because such articulation changes the conclusion about using a particular model. Second, the current ApAr approach does not provide uncertainty estimation, but there is an ongoing effort to provide confidence bounds, to better enable empirical comparison of models. Third, we have explored the application of our approach to binary classification problems only and have not studied multiclass problems. Fourth, the current approach holds fixed utility tradeoffs. Fifth, the current approach focuses on utility. Health system decision-makers may prefer a metric that includes financial considerations, such as net monetary benefit, as DCA does. Future work will aim to include ranges of tradeoffs and alternative utility formulations in the model evaluation. In establishing a clinically relevant performance characteristic, model construction also demands coherence between asymmetric costs in the model-development loss function and the asymmetric cost used in ApAr evaluation. Future work will investigate the role of asymmetric cost in different ML modeling approaches. We also aim to incorporate more state-of-the-art deep learning models to understand how ApAr and cost asymmetry might be of value in image analysis for instance. Lastly, our approach is not yet computationally optimized.

## Conclusions

The Applicability Area is a novel metric that addresses the next phase of SMLM adoption: Not only does it perform well, statistically, but will it be useful in the decisional contexts for which it is intended. Given the considerable number of such models being developed, attention to this adoption phase is crucial. We hope that our principle-based metric will be of use to such decision-makers.

## Data Availability

All data produced are available upon reasonable request to the authors.

https://github.com/StarLiu1/ApplicabilityArea-ApAr

## Acknowledgment

We would like to give special thanks to Dr. Hadi H.K. Kharrazi, Dr. Michael C. Higgins, and Dr. Stephen M. Downs.

